# Multi-omic Biomarkers Distinguish Rheumatoid Arthritis in Discordant Monozygotic Twins

**DOI:** 10.1101/2024.12.30.24319783

**Authors:** Rebecca B. Blank, Kevin Bu, Weixi Chen, Ian Cunningham, Jeremy Sokolove, Lauren Lahey, Adriana Heguy, Rhina Medina, Carles Ubeda, Renuka R. Nayak, Jiyuan Hu, Adam Cantor, Jakleen Lee, Jose C. Clemente, Jose U. Scher

## Abstract

**Background:** Although genetic factors have been identified in the pathogenesis of rheumatoid arthritis (RA), the concordance rate in monozygotic (MZ) twins is low, suggesting that other features contribute to disease development. Further, the relative contribution of such non-genetic elements in identical twins have not been characterized. Here, we aimed to measure differentiating host and microbial biomarkers of RA by studying MZ twins discordant for disease using a multi-omics approach.

**Methods:** Eight pairs of MZ twins discordant for RA (n=16) were enrolled. Gut microbiome was assessed using shotgun metagenomic sequencing. Autoantibodies, cytokines, and other plasma proteins were measured in both plasma and feces. Levels of short and medium-chain fatty acids from serum and feces were quantified using gas chromatography mass spectrometry (GC-MS).

**Results:** While overall microbiome diversity and composition did not significantly differ between twins, we observed a decrease in *Blautia faecis* in affected twins. Affected twins had higher concentrations of both fecal and plasma citrullinated and non-citrullinated autoantibodies, as well as significantly lower concentrations of fecal butyrate and propionate.

**Conclusion:** Multi-omics biomarkers differentiate MZ twins discordant for RA. *Blautia faecis*, which is associated with reduced inflammatory cytokine expression, was decreased in RA twins. Similarly, short-chain fatty acids, known to have immune modulatory effects, were decreased in affected twins, suggesting further bi-directional interactions between inflammation at the gut barrier and disease state. If confirmed in other cohorts, exhaustive multi-omics approaches may improve our understanding of RA pathogenesis and potentially contribute to novel diagnostics and co-adjuvant therapies.

## Introduction

Although a number of genetic factors, including susceptibility alleles, have been identified in the pathogenesis of rheumatoid arthritis (RA), the concordance rate in monozygotic (MZ) twins is only 12 to 15%^1,2^. This observation suggests that other factors also contribute to disease onset and perpetuation. Among those, the microbiome, autoantibodies, metabolome and proteome have all been associated with RA.

The gut microbiome has been shown to be associated with inflammatory arthritis in various murine models^3–5^, and different studies have implicated microbial taxa in the pathogenesis of human RA, including periodontal microbes *Prevotella gingivalis*^6,7^, *Lactobacillus salivarius*^8^ (present in both saliva and gut) and gut microbes *Prevotella copri*^9,3,10^ and *Subdoligranulum*^11^, among others^12^. *Prevotella copri* is more abundant in new-onset RA and appears to stimulate strain-specific immune responses in patients ^9,10^. In fact, some of these same microbes have been implicated in the induction of RA-associated pathogenic autoantibodies which can be observed in both pre-RA^7^ and symptomatic RA patients^6,8,10,11^. In turn, these microbes can activate innate-, B– and T-cell responses that then produce pro-inflammatory cytokines, such as IL-17^3,4,10^, IL-1, IL-6, TNF-α, and others^12^. Conversely, gut microbial fermentation byproducts, such as short chain fatty acids (SCFAs), have been shown to induce a tolerogenic immune response^13^, ameliorate inflammatory arthritis in murine models^14,15^ and found to be decreased in serum concentrations in pre-RA individuals^16^. While several studies have investigated the contribution of each of these factors to RA in isolation, few have integrated different assays to characterize the molecular landscape of immune-mediated inflammatory disease^17–19^. Critically, no study to date has interrogated differentiating features using a multi-omics approach while controlling for host genetics. Here, we combined data derived from microbiome, autoantibodies, metabolome, and circulating plasma proteins in a relatively small albeit unique cohort of MZ twins discordant for RA to identify disease signatures.

## Methods

### Participants

Monozygotic (MZ) twin pairs discordant for RA were recruited from the population-based Mid-Atlantic Twin Registry (MATR). Eight pairs of MZ twins were enrolled in the study. Diagnosis of RA was obtained through patient medical records. Upon enrollment, demographics, detailed medical history, including review of past and present medication were collected. In addition, MD-HAQ and swollen joint counts (SJC) and tender joint counts (TJC) were estimated. Stool, serum, and plasma samples were collected at time of enrollment.

### Patient and public Involvement

As this was a cross-sectional study, involving only one time-point, we did not involve patients or the public in the study design.

### Metagenomics

Stool samples underwent bacterial DNA extraction and shotgun sequencing to characterize the composition and function of the gut microbiome as previously described^20^. Metagenomic analysis of sequencing data was performed using Metaphlan 4^21^. Pathway analysis was conducted using Humann 3^22^. Diversity metrics and visualizations were done in QIIME2 V2020.8.0. Shannon Diversity was used to estimate alpha diversity, while Bray-Curtis was used for beta diversity. The Bray-Curtis distance matrix was used as input for principal coordinate analysis (PCoA). Functional annotation was done using Humann3 and the BriteKO hierarchy.

### Multiplex autoantibody assays

Autoantibody concentration was determined in stool and plasma using multiplex flow cytometry assay as previously described ^23,24^. Briefly, a panel of 103 citrullinated and non-citrullinated self-protein antigens were conjugated to fluorescent beads using the Bio-Plex multiplex assay platform (BIO-Rad Laboratories, Hercules, CA, USA) and analyzed using the Luminex 200 (Luminex, Austin, TX, USA). Pooled beads were mixed with either plasma or stool samples and incubated at room temperature. After washing, the sample-dyed bead mixture was incubated with anti-human IgG antibody conjugated to phycoerythrin (PE) at room temperature. After second wash, the bead mixture was passed through the laser detector, Luminex 200, that identified the beads based on fluorescence of the dyes. Quantity of autoantibody bound to each bead was determined by PE fluorescence and expressed as mean fluorescence intensity (MFI). Serum anti-CCP antibody was measured by QuantaLite CCP 3.1 IgG/IgA ELISA (INOVA Diagnostics, Fairfax, VA, USA).

### Analysis of plasma cytokines and proteins

Plasma analytes were measured by proximity-extension assay (PEA) (OLINK technology, Bioxpedia Aarhus, Denmark) using a 92 inflammation-associated plasma protein biomarker panel. PEA was performed at the Human Immune Monitoring Center, Icahn School of Medicine, NY, NY, USA. PEA allows for detection of very low concentrations of plasma analytes and the final concentration is given in Normalized Protein eXpression (NPX)^25^ units, which is an arbitrary unit on a log2 scale. Of the 92 analytes in the panel, six analytes were removed from analysis because at least 33% of the sample measurements were below the limit of detection.

### Metabolomics

Fecal samples were collected, stored on freezer packs and shipped to the Scher lab within 24 hours of collection and immediately stored at –80°C as previously described^20^. Fecal short– and medium-chain fatty acids (SCFA, MCFA) measurements were performed at the University of Michigan Metabolomics Core (Michigan Medicine Biomedical Research Facilities, Ann Arbor, MI, USA). Briefly, SCFA analysis was performed by GC-MS on an Agilent 69890N GC-5973 MS detector as previously described^26^. MCFA analysis was performed by RPLC-MS on an Agilent 1290 Infinity II / 6545 qTOF MS system with the JetStream Ionization (ESI) source (Agilent Technologies, Inc., Santa Clara, CA USA) using the Waters Acquity HSS T3 1.8 µm 50 mM column (Waters Corporation, Milford, MA). Each sample was analyzed twice, once in positive and once in negative ion mode. Mobile phase A was 100% water with 0.1% formic acid and mobile phase B was 100% methanol with 0.1% formic acid. The gradient for both positive and negative ion modes was as follows:.2% B (0 min), 75% B (20 min), 98%B (22 min), 98%B (30 min), 2% B (30.1 min) was used. The column was then reconditioned for 7 min with 2%B before moving to the next injection The flow rate was 0.46 mL/min and the column temperature was 40°C. The injection volume for positive and negative mode was 5 µL and 8 µL, respectively. Source parameters were: drying gas temperature 350°C, drying gas flow rate 10 L/min, nebulizer pressure 30 psig, sheath gas temp 350°C and flow 11 l/min, and capillary voltage 3500V, with internal reference mass correction. Data analysis of SCFA and MCFA were processed using MassHunter Quantitative analysis version B.07.00. SCFAs and MCFAs were normalized to the nearest isotope labeled internal standard and quantitated using 2 replicated injections of 6 standards to create a linear calibration curve with accuracy better than 80% for each standard. Other compounds in the analysis were normalized to the nearest internal standard, and the peak areas were used for differential analysis between groups. Samples were normalized to wet sample weight after quantification.

Serum samples were collected, centrifuged at 1700g for 10 minutes and immediately stored at – 80° C. Frozen serum sample aliquots were shipped on dry ice for SCFA metabolomics profiling by Metabolon Inc. (Morrisville, NC), as previously described^27^, using the quantitative SCFA panel. Quantitative analysis was performed by LC-MS/MS on an Agilent 1290 UHPLC/Sciex QTrap 5500 instrument.

### Statistical Analyses

Association of clinical metrics (e.g. age, sex, ethnicity, swollen and tender joint counts) with disease grouping was assessed using Fisher’s Exact Test for dichotomous variables and paired t-tests for continuous variables. Analysis of inflammatory cytokines, fatty acids, and citrullinated peptides was performed using paired t-tests. Taxonomic and functional metagenomic data were analyzed using Mann-Whitney U, Kruskal-Wallis, and Wilcoxon Signed Rank tests. Kruskal-Wallis and permutational multivariate analysis of variance (PERMANOVA) with 999 permutations were used to compute differences in beta diversity.

Comparison of microbiome diversity distances between three groups (intra-twin, inter-RA and inter-unaffected) was done as follows. For alpha diversity, the intra-twin distances were obtained by computing the absolute value of the differences in Shannon Entropy for each of the twin pairs (|α_Unaffected_ – α_RA_|). For the inter-RA distances, the absolute value of the difference in Shannon Entropy was computed for each pair of RA-affected twins, i.e. for all (8 choose 2) = 28 pairs. Analogous distances were computed for the inter-unaffected group.

Beta diversity distances were computed in a similar fashion. The intra-twin distances consist of the 8 Bray-Curtis distances between unaffected and affected twins, i.e. one per twin pair. The inter-RA differences consist of aggregating all (8 choose 2) = 28 distances between each pair of individuals with RA, and an analogous calculation was performed for the inter-unaffected differences. The distances of these three groups (intra-twin, inter-RA, and inter-unaffected) were then compared using the Kruskal-Wallis test. Correlations of bacterial diversity (Shannon entropy for alpha diversity or Bray-Curtis intra-twin for beta diversity) with SJC, TJC, CRP, and DAS28-CRP were performed using Spearman correlation. Cohen’s D effect size was computed using the method previously described^28^.

Statistical analyses (paired and unpaired t-tests, Wilcoxon, Mann-Whitney U, Cohen’s D), and plotting were performed using Python 3.7 and required libraries (numpy, pandas, matplotlib, seaborn), and R 4.2.1 and libraries (phyloseq, reshape2, vegan, ade4, PMCMRplus, ggplot2, tidyverse, dpylr, tidyr, scales, ComplexHeatMap). Unless otherwise specified, functions, plotting, and testing were performed with default parameters.

## Results

Eight monozygotic twin pairs discordant for RA were enrolled with a mean age of 56.3 years and mean disease duration of 7.0 years (**Table 1**). The majority were female (87.5%), and all were non-hispanic white. RA twins had moderate disease activity with a mean disease activity score-C-reactive protein (DAS 28-CRP) of 4.02. Their mean tender joint count (TJC) was 10.4 and mean swollen joint count (SJC) was 6.25. Both affected and unaffected twins had a large range of serum anti-cyclic citrullinated peptide (CCP) antibody levels as quantified by CCP 3.1 IgG/IgA ELISA, with no significant difference between affected and unaffected twins. Most RA twins (75%) were currently or had previously taken methotrexate (MTX) and 62.5% were currently taking MTX at time of the study. 62.5% of RA twins were being treated with or had previous exposure to a biologic agent (**Table 1**); four RA twins had exposure to TNF inhibitors and one had had exposure to multiple TNF inhibitors and abatacept and was on rituximab at the time of enrollment. None had taken or were currently taking a JAK inhibitor.

**Table 1:**
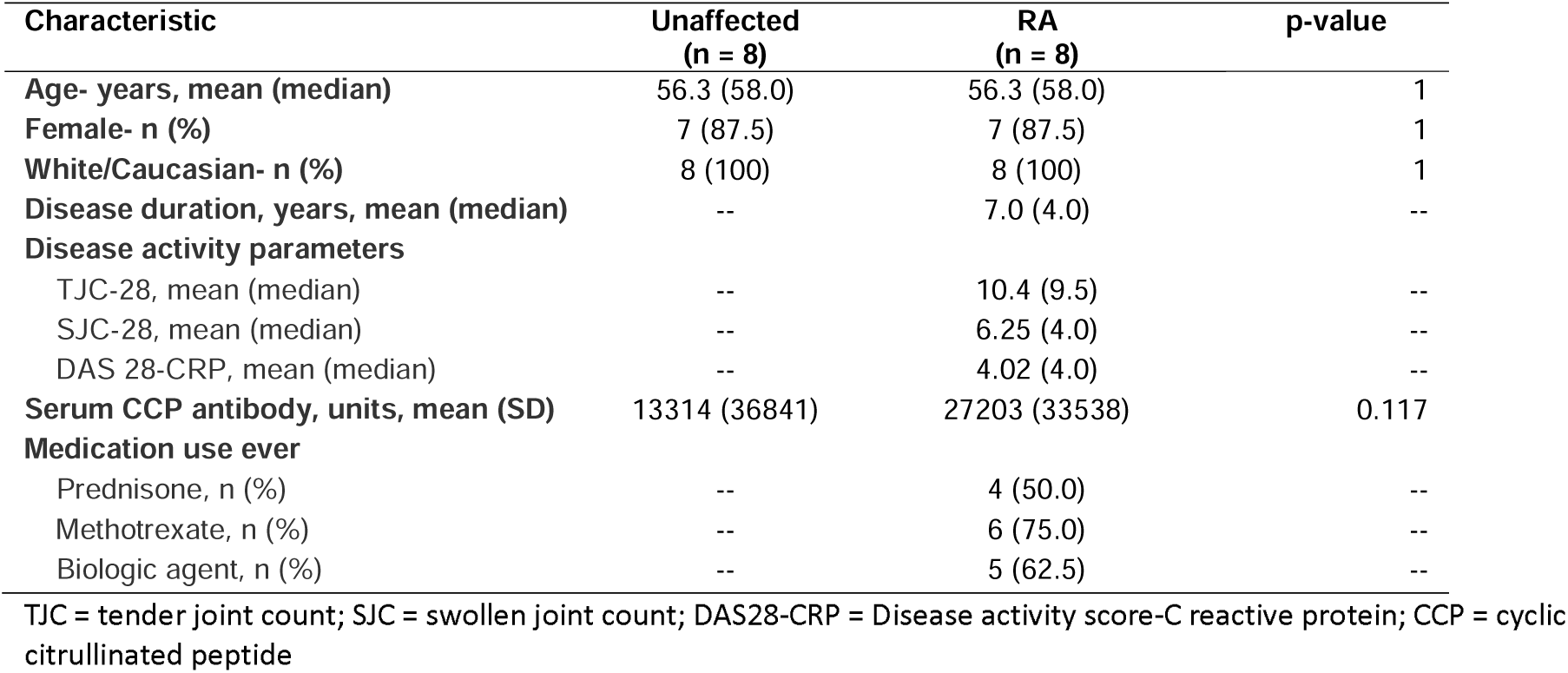
Cohort demographics and clinical information.

| Characteristic | Unaffected<br>(n = 8) | RA<br>(n = 8) | p-value |
| --- | --- | --- | --- |
| Age- years, mean (median) | 56.3 (58.0) | 56.3 (58.0) | 1 |
| Female- n (%) | 7 (87.5) | 7 (87.5) | 1 |
| White/Caucasian- n (%) | 8 (100) | 8 (100) | 1 |
| Disease duration, years, mean (median) | -- | 7.0 (4.0) | -- |
| **Disease activity parameters** |  |  |  |
| TJC-28, mean (median) | -- | 10.4 (9.5) | -- |
| SJC-28, mean (median) | -- | 6.25 (4.0) | -- |
| DAS 28-CRP, mean (median) | -- | 4.02 (4.0) | -- |
| **Serum CCP antibody, units, mean (SD)** | 13314 (36841) | 27203 (33538) | 0.117 |
| **Medication use ever** |  |  |  |
| Prednisone, n (%) | -- | 4 (50.0) | -- |
| Methotrexate, n (%) | -- | 6 (75.0) | -- |
| Biologic agent, n (%) | -- | 5 (62.5) | -- |

TJC = tender joint count; SJC = swollen joint count; DAS28-CRP = Disease activity score-C reactive protein; CCP = cyclic citrullinated peptide

### Gut microbiome of RA and unaffected twins

Using shotgun sequencing, we first compared the gut microbiome of RA affected and unaffected twins. Alpha and beta diversity were not significantly different between twins (Mann-Whitney U p=0.95 and PERMANOVA p=0.80, respectively; **Figures 1A-B**). We then analyzed the gut microbiome of individuals with a shared diagnosis versus individuals with a shared genetic background. For alpha diversity, we compared the absolute differences in Shannon Entropy for all 8 twin pairs (unaffected minus affected) with the absolute differences in Shannon Entropy for all pairs of individuals in the RA-affected and unaffected groups i.e. (8 choose 2) = 28 distances for each group separately. Similarly, for beta diversity, the Bray Curtis distances were computed for the 8 twin pairs and compared to the 28 distances between all pairs of RA-affected and the 28 distances between all pairs of unaffected individuals. There were no significant differences between intra-twin and intra-disease distances in overall species diversity using Shannon Entropy (Kruskal-Wallis p=0.67, **Supplementary Figure 1A**). In beta diversity, twin pairs tended to be more similar to each other than unrelated individuals with a shared diagnosis, although not significantly (Kruskal-Wallis p=0.15**, Supplementary Figure 1B**).

**Figure 1.**
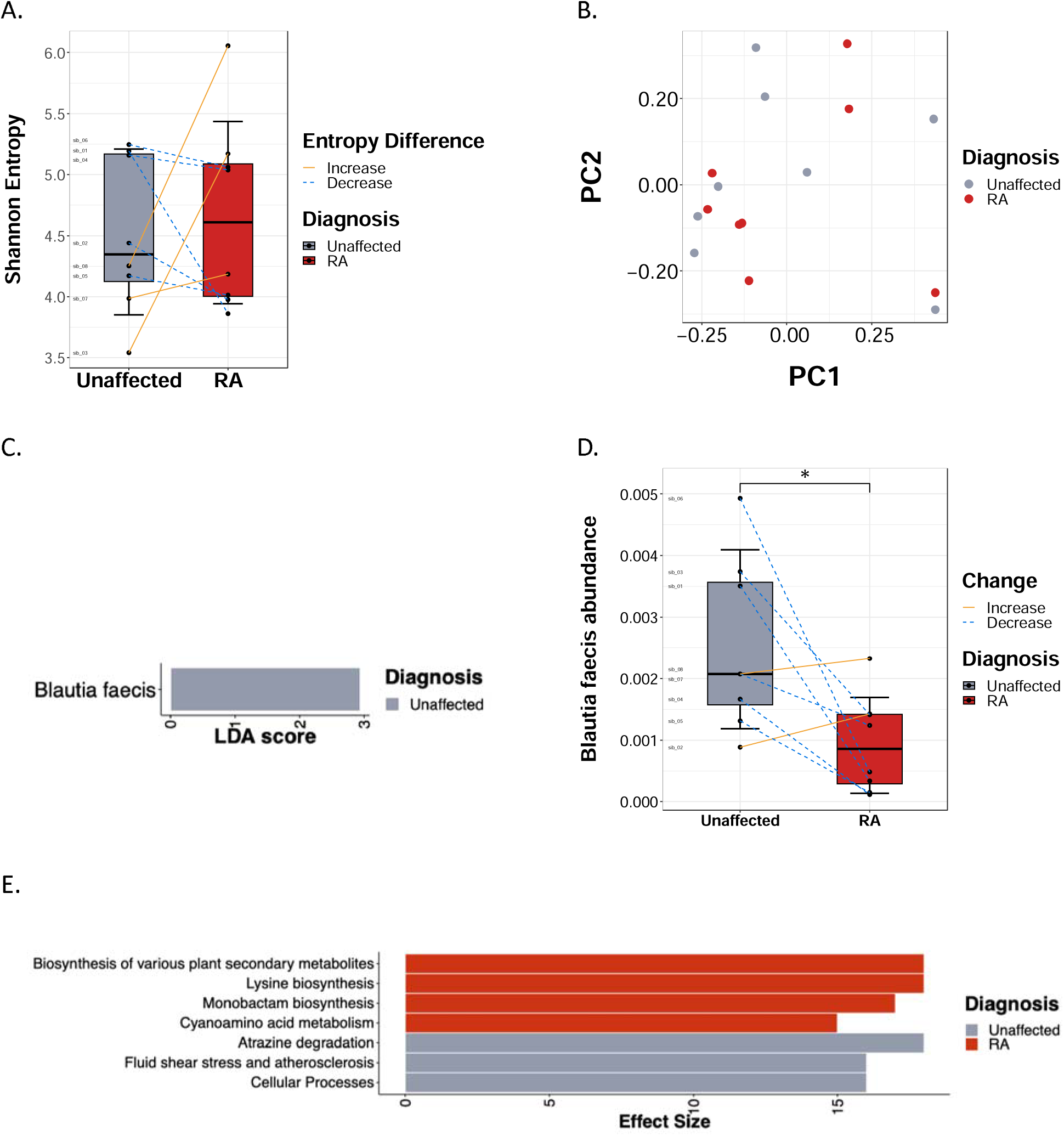
Gut bacterial composition and predicted functional pathways in RA and unaffected twins. (A) Shannon alpha diversity in participants, with box plots representing median and interquartile range. Lines are drawn between MZ pairs. (B) Principal coordinate analysis (PCoA) of Bray-Curtis beta diversity between groups. (C) Linear discriminant analysis Effect Size (LEfSe) showing differential species in RA twins compared to unaffected twins. (D) Boxplot showing distribution of *B. faecis* abundance in unaffected and RA twins. (E) Barplot of Wilcoxon effect sizes for differential pathways. *p<0.05, **p<0.01.

When analyzing individual taxa, Linear discriminant analysis Effect Size (LEfSe) found a decrease of *Blautia faecis* in RA twins (Wilcoxon p=0.04) (**Figures 1C-D**). Analysis of functional features also identified 7 pathways that were differentially abundant between RA and unaffected twins, including lysine metabolism, cyanoamino acid metabolism, and monobactam biosynthesis, which were more abundant in RA (**Figure 1E, Supplementary Figure 2**).

We then investigated the effect of MTX use on microbiome composition. Current treatment with MTX was not associated with significant differences in alpha (Mann-Whitney U p=0.39, **Supplementary Figure 3A**) or beta diversity (Mann-Whitney U p=0.57, **Supplementary Figure 3B**). Alpha diversity was not significantly associated with disease activity, as measured by swollen joint count (SJC) (Spearman rho=0.11, p=0.8), TJC (Spearman rho=0.36, p=0.39), C-reactive protein (CRP) (Spearman rho=-0.10, p=0.82) and DAS28-CRP (Spearman rho=0.12, p=0.78) (**Supplementary Figure 4 A-D**). Furthermore, we found no association involving these four clinical measures when correlating these metrics for the RA-affected twin with the intra-twin beta diversity (p=n.s.)

### Short-chain fatty acids are decreased in affected twins

We next performed targeted metabolomics to determine whether there were differences in fatty acid concentration between affected and unaffected twins. Compared to their unaffected siblings, RA twins had significantly lower concentrations of fecal butyrate (p=0.03), propionate (p=0.03), and acetate (p=0.05) and a significantly higher concentration of fecal valerate (p=0.04) (paired t-test, **Figure 2A-D**). There was no significant difference in serum SCFAs between the two groups although we observed a trend towards higher concentrations of SCFA in the unaffected twins (paired t-test; p=0.41 for 2-methylbutyric acid, p=0.31 for isobutyric acid, p=0.67 for acetic acid, and p=0.15 for hexanoic acid in **Supplementary Figure 5**).

**Figure 2.**
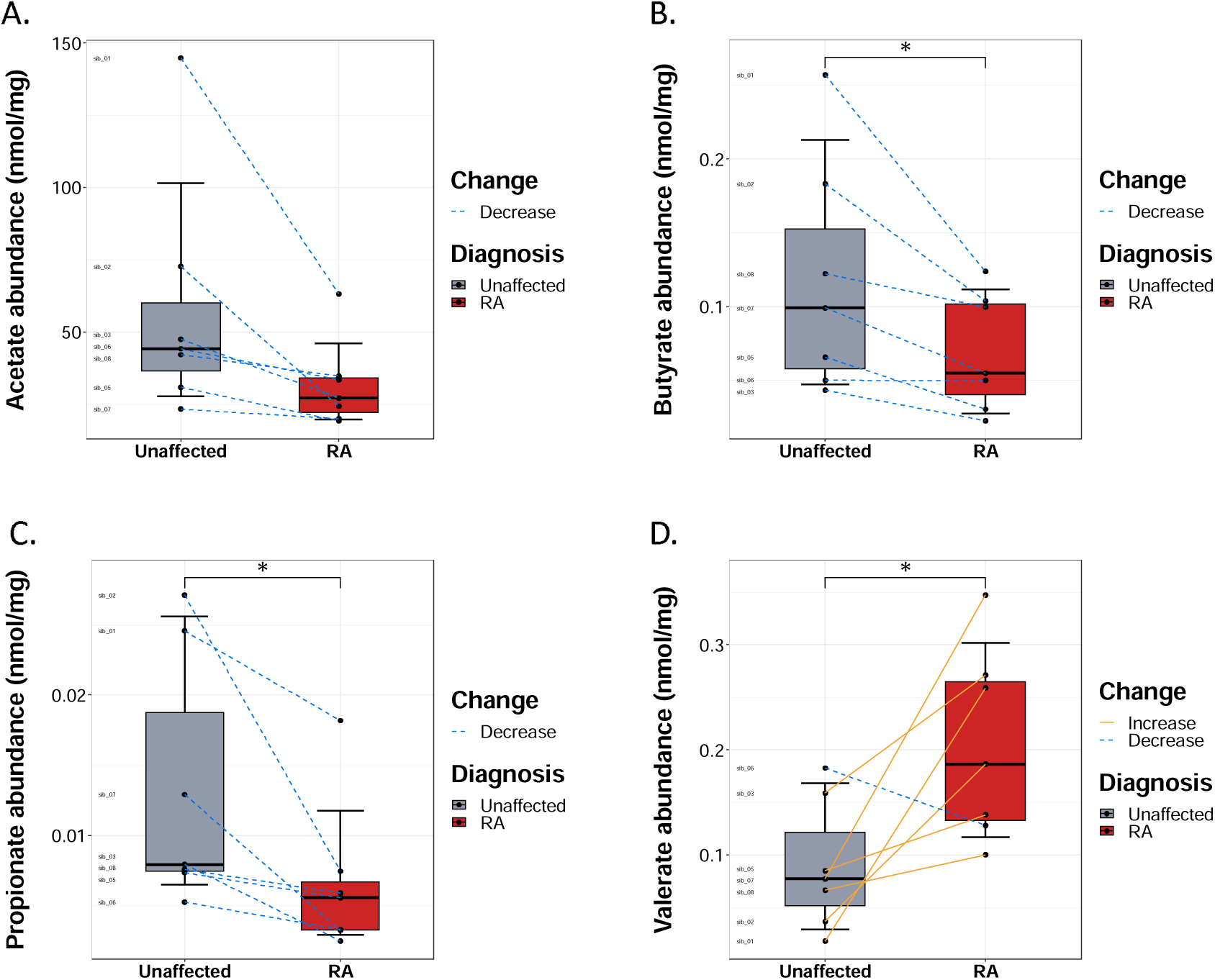
Comparison of fecal short chain fatty acid (SCFA) concentrations between unaffected participants and RA twins. (A) RA twins have lower fecal concentration of acetate. (B) Butyrate fecal levels are depleted in RA twins. (C) Propionate fecal concentration is lower in affected twins. (D) Valerate fecal levels are higher in RA twins. Lines are drawn between MZ pairs. P-values calculated with paired t test; *p<0.05.

### RA twins have elevated fecal and plasma autoantibodies

We next explored whether autoantibodies considered to be pathogenic in RA were differentially expressed in twin pairs. Using multiplex bead assay, we measured 103 fecal and plasma autoantibodies (listed in **Supplementary Table 1**) against either citrullinated (ACPA) or non-citrullinated native proteins and found one fecal autoantibody and 14 plasma autoantibodies that were significantly elevated in RA twins compared to their unaffected siblings (**Figure 3 A-B and Supplementary Table 2**). Among the 15 significantly elevated autoantibodies, we noted autoantibodies to enolase, fibrinogen, vimentin and others.

**Figure 3.**
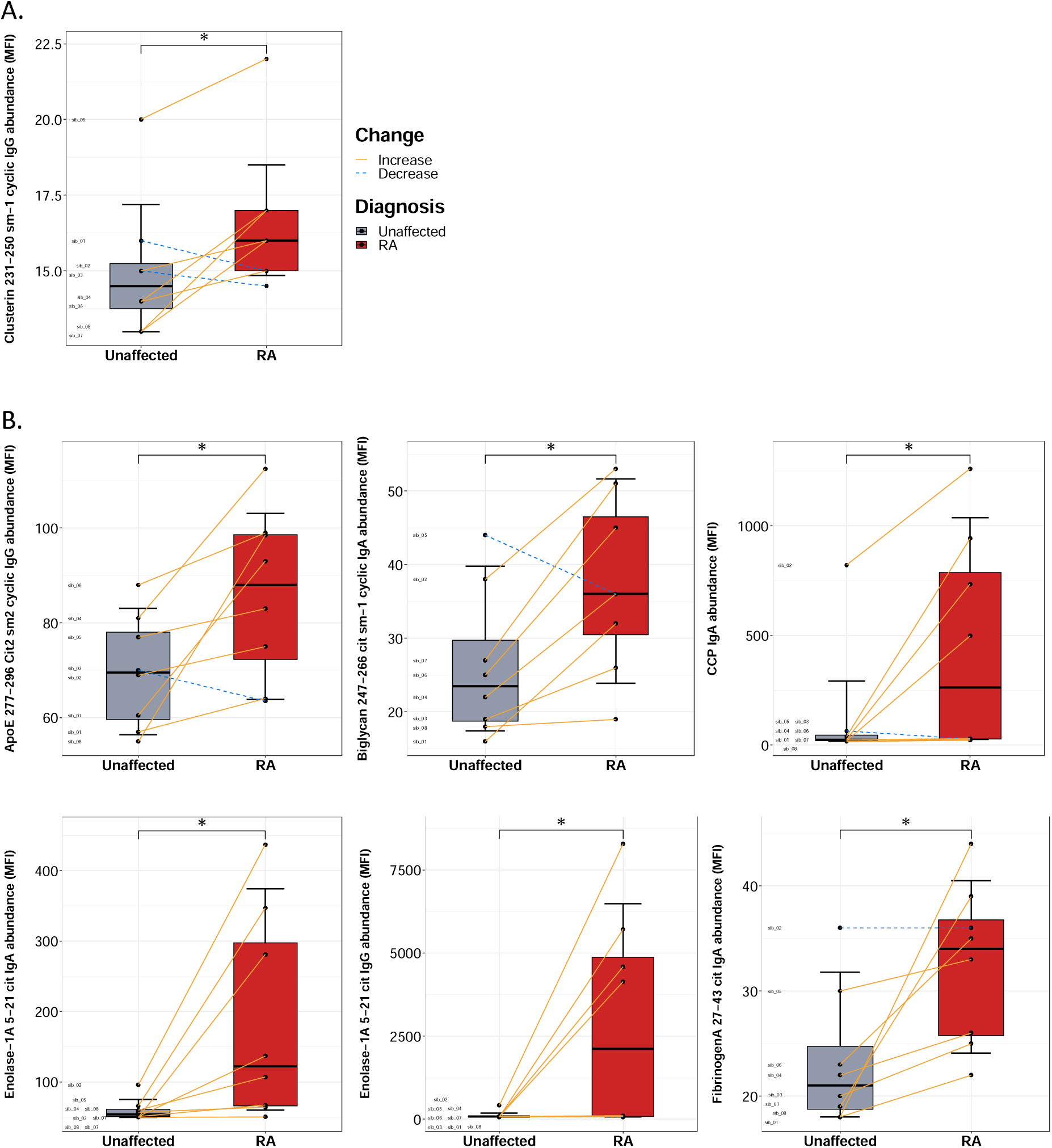
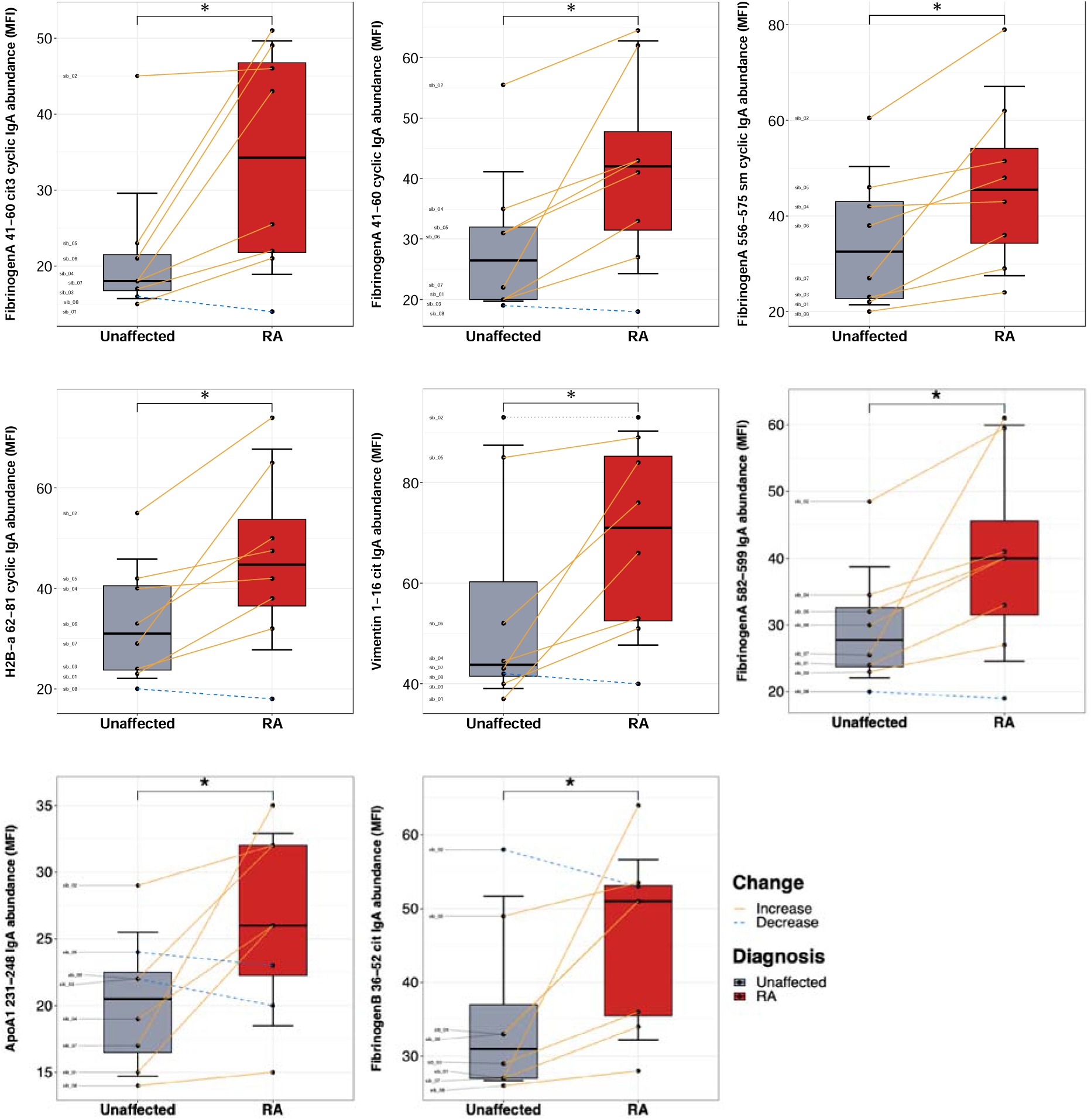
Comparison of citrullinated and non-citrullinated fecal and plasma autoantibody concentration between unaffected and RA twins as measured by mean fluorescence intensity (MFI). Fecal autoantibody boxplots (A) and plasma autoantibody boxplots (B) are shown. Box plots represent median and interquartile range. Lines are drawn between MZ pairs. P-values calculated with paired t test; *p<0.05.

### Elevated circulating inflammatory proteins in affected twins

Using a highly sensitive proximity extension assay (PEA) to detect low levels of protein in the plasma, we measured 92 analytes using the OLINK Target 96 Inflammation Panel. We found that five were significantly elevated in RA twin plasma compared to unaffected twins (**Figure 4 A-E**) whereas none were significantly elevated in unaffected twins. IL-1α, the chemokines CXCL1, CCL23, CCL25, and matrix metalloproteinase MMP-10 were all elevated in RA twins (paired t-test; p=0.003, p=0.007, p=0.026, p=0.041, and p=0.041, respectively).

**Figure 4.**
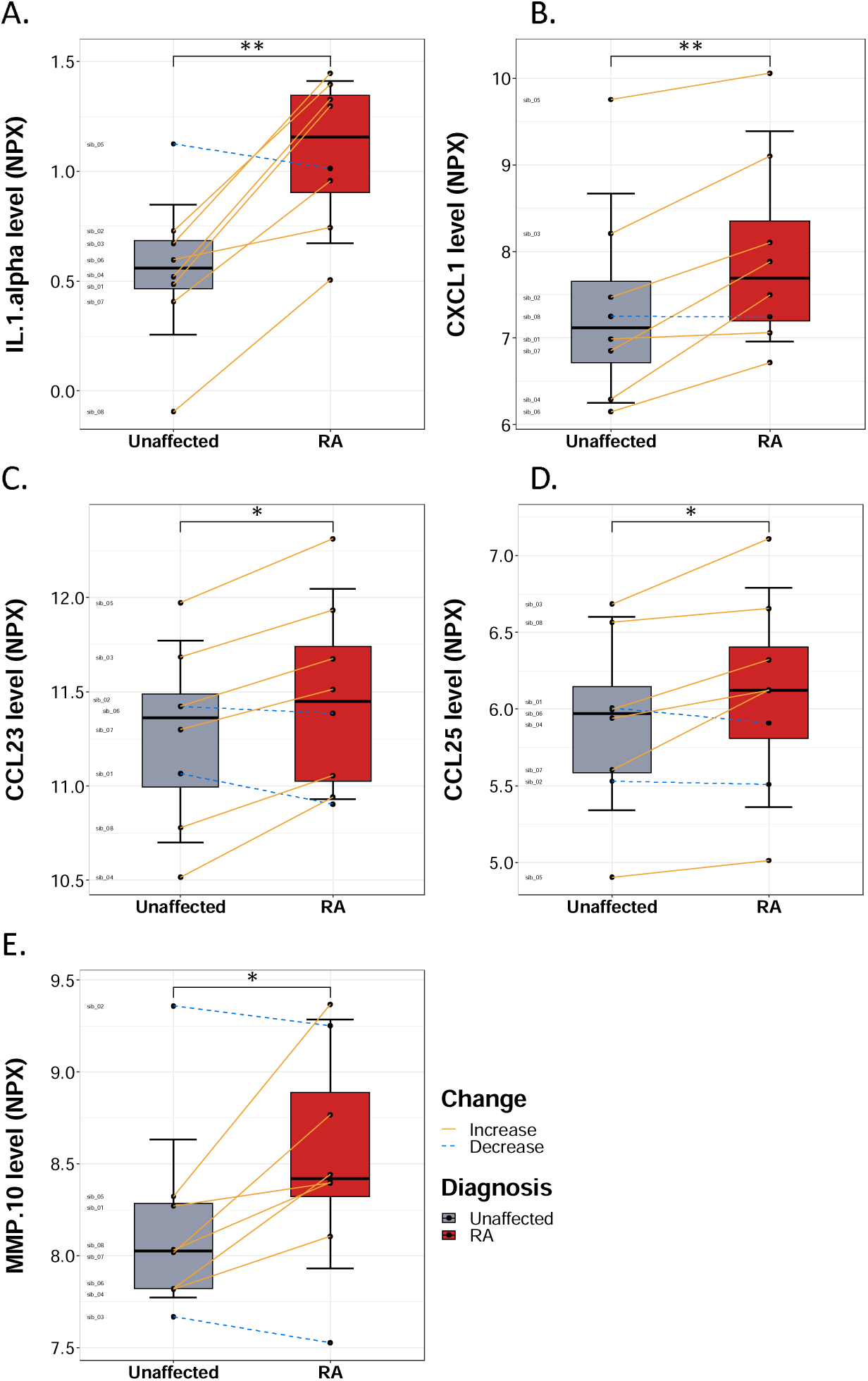
Levels of circulating cytokines and other proteins in RA and unaffected twins. Proximity extension analysis of 92 inflammation-related plasma proteins revealed significant elevation of (A) IL-1α, (B) CXCL1, (C) CCL23, (D) CCL25, and (E) MMP-10 in RA versus unaffected twin. Y axes represent normalized expression values (NPX) in log2 scale. Box plots represent median and interquartile range. Lines are drawn between MZ pairs. P-values calculated with paired t test; *p<0.05, **p<0.01.

### B. faecis, monobactam biosynthesis, and cyanoamino acid metabolism are associated with inflammatory markers

We conducted Spearman correlation analysis between taxa and functional pathways against cytokines, fatty acids, and autoantibody data in the RA and unaffected twins separately, focusing on the differential taxa (*B. faecis*) and 7 differential pathways. We found no significant correlations involving *B. faecis* and 34 involving differential pathways in the RA group (two of which also involved differential inflammatory markers), and 30 significant correlations involving *B. faecis* and 48 involving differential pathways in the unaffected twins (all p<0.05), after filtering out false positive results using CUTIE^29^. Out of the 30 significant correlations involving *B. faecis* in the unaffected twins, two were with cytokines and 28 were with fecal ACPAs. *B. faecis* was positively correlated with FMS-like tyrosine kinase 3 ligand (Flt3L) in unaffected twins (rho=0.90, p=0.002) and negatively correlated with stem cell factor (SCF) (rho, –0.98, p=0.00003; **Supplemental Figure 6 A-B**). Additionally, all but one of the remaining 28 significant correlations involving *B. faecis* and fecal autoantibodies were negative (rho < 0). We also examined the correlations between pathways and inflammatory markers and found that two of the 82 correlations involving differential pathways also involved differentially expressed inflammatory markers. Both correlations were with CXCL1; in the RA twins, CXCL1 was positively associated with cyanoamino acid metabolism (rho=0.88, p=0.004) and monobactam synthesis (rho=0.86; p=0.007; **Supplementary Figure 7A-B**).

### Comparison of Effect Sizes Across –Omics Data Types

Lastly, we compared the effect size of the top three differential features of each data type using the paired Cohen’s D statistic (**Figure 5**). Among the metagenomic pathways, biosynthesis of plant secondary metabolites, lysine biosynthesis, and atrazine degradation exhibited the largest effect sizes, while among the inflammatory markers, IL1-α exhibited the greatest Cohen’s D (larger than 1.5), followed by CXCL1 and MMP 10. The SCFAs butyrate, propionate, and valerate had large effect sizes, with Cohen’s D approaching 1, as did the ACPA plasma Fibrinogen A 27-43 cit IgA, and bacteria *Blautia faecis*, also with Cohen’s D approaching 1.

**Figure 5.**
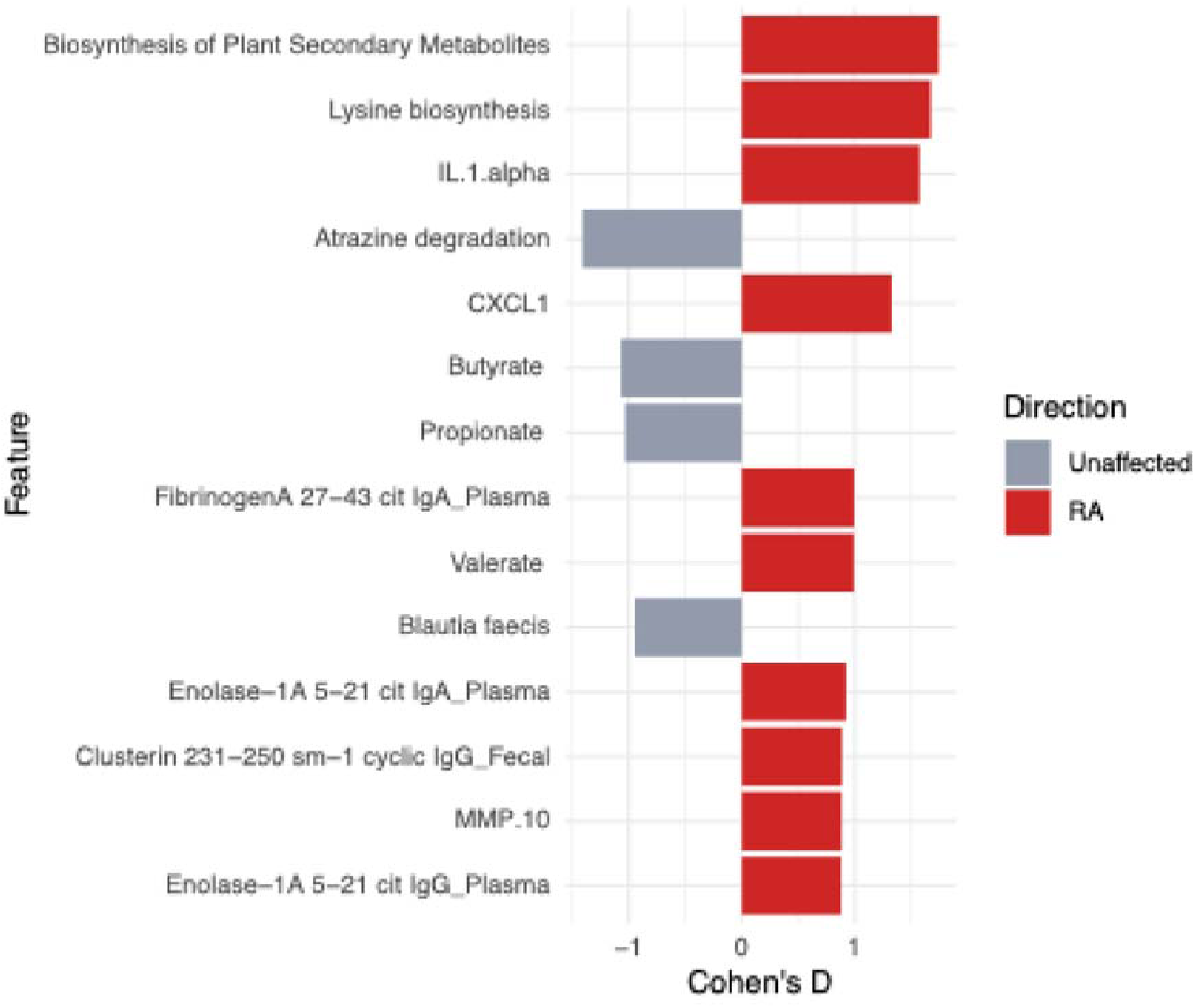
Cohen’s D (paired) effect sizes for top differential features of each datatype along with directionality of enrichment.

## Discussion

While studies on twins have been previously conducted to determine concordance rates in RA, ours is the first to perform an integrated multi-omics investigation of microbial, immune, and metabolic features that differentiate monozygotic twins discordant for RA. Two prior studies have used a multi-omics approach to study RA, although limited to studying gut microbiome and metabolomic differences between RA patients and unrelated (non-twins) healthy controls^18,19^. Another recent study assessed the differences in gut microbiome, colonic transcriptome and peripheral blood immune profiles between patients with psoriasis and healthy controls^17^. While we found no overall differences in microbial alpha or beta diversity between RA and their unaffected twins, we identified one species, *Blautia faecis,* that was differentially abundant. *Blautia faecis*, which produces lactate and acetic acid and reduces expression of inflammatory cytokines such as TNFα and IL-8^30,31^, was enriched in unaffected twins. Flt3L, which has been implicated in both inflammatory^32^ and regulatory^33,34^ processes in RA, was positively correlated with *B. faecis* abundance in unaffected twins, suggesting that *B. faecis* may modulate a regulatory process via its association with Flt3L that could contribute to protection against RA. This observation was further confirmed by the negative correlation between fecal autoantibodies and *B. faecis* in the unaffected twins. Additionally, *Blautia spp*. are butyrate and propionate producers^30^, which may explain the increased levels of fecal SCFA observed in the unaffected twins.

The lack of differences in microbiome diversity between affected and unaffected twins is not unexpected, given that the RA twins had been treated or were undergoing treatment at time of sample collection. In fact, the gut microbiome of untreated RA and healthy controls are significantly different^9^ but those differences are partially reduced after treatment^5,8,35,19^. Effective RA treatment may also contribute to our observation that microbial composition was more similar between intra-pair MZ twins than between unrelated individuals of the same disease state, either unaffected or RA.

The anti-inflammatory properties of gut microbial fermentation products such as SCFAs have been well-studied in both murine models of RA and humans^14,15,36^ and play a protective role against disease progression in murine models. SCFA levels such as propionate, remained low in early RA patients both before and after MTX treatment^19^, suggesting that, in our cohort, reduced SCFA in RA twins was more likely due to the disease state and not an effect of treatment regimen. Additionally, elevated serum levels of butyrate and acetate have been associated with non-progression to inflammatory arthritis in pre-RA individuals^16^ and in psoriatic arthritis^37^. We found that SCFAs are elevated in the feces of unaffected twins, further confirming that SCFAs may play a protective role in individuals that are genetically predisposed to developing RA.

Although most RA twins were being treated with either csDMARDs and/or bDMARDs at the time of enrollment, we found that RA twins had elevated plasma levels of inflammatory cytokines and chemokines nonetheless. Interestingly, some of the significantly overexpressed proteins, such as IL-1α, CCL23, CCL25, CXCL1, and matrix metalloproteinases, have also been recognized as potential biomarkers of RA in previous studies ^38–43^ and are known to play a role in synovial inflammation^41,44^, tissue remodeling^43^ and osteoclastogenesis^40,45^. RA twins had higher levels of citrullinated and non-citrullinated plasma and fecal autoantibodies than their unaffected siblings. These included autoantibodies to enolase, fibrinogen, filaggrin, and vimentin, all previously implicated in RA pathogenesis^23,24^. It is unclear whether dysregulation at the mucosal surfaces, such as the gut, drive autoantibody expansion and differentiation or whether development of RA itself drives microbial dysregulation. Genetically, both unaffected and RA twins are equally predisposed to developing pathogenic autoantibodies associated with RA. Therefore, it is plausible that dysregulation of the gut microbiome at the intestinal barrier of affected twins prior to RA development could lead to modifications of self-proteins that subsequently trigger the production of pathogenic autoantibodies, as in the case of periodontal disease^6,7^. Further, unaffected twins without microbial dysbiosis may not be primed to develop pathogenic autoantibodies and are therefore more likely to be spared from developing RA, a situation similar to what has been shown in pre-RA^11^ and early RA individuals^9^.

We recognize several limitations to our study. Although of unique value, the size of our cohort is relatively small and a larger set of discordant twins will be needed to validate our findings. One unaffected twin was eventually diagnosed with RA after conclusion of the study and may have confounded some analyses. Additionally, most RA twins had been on DMARD treatment at time of enrollment, and the mean duration of their disease was 7 years. This represents a confounder when measuring differences between molecular features and would require future studies to include new onset RA twins to compare against their discordant twins. Further, our study is also cross-sectional in nature, which prevents us from investigating how disease pathogenesis and treatment impact results. Despite these limitations, we were able to identify several differentiating features between RA and their unaffected twins. Regardless of treatment status, RA twins had higher levels of circulating inflammatory cytokines and chemokines and lower levels of tolerogenic SCFAs. They had higher levels of circulating pathogenic autoantibodies and, importantly, they were found to have a significantly lower abundance of *B. faecis*, a species that is implicated in a reduced proinflammatory response. We note that this is the first study that uses a multi-omics approach to investigate microbial, metabolomic and molecular biomarkers in twins discordant for rheumatoid arthritis. The unique setting of our cohort thus provides a novel avenue to investigate RA pathogenesis while controlling for genetic predisposition and highlights the need to integrate fecal metabolomics and host molecular features to microbiome studies.

## Data Availability

All data produced in the present study are available upon reasonable request to the authors.

## Acknowledgements

The project was facilitated by the Mid-Atlantic Twin Registry (MATR) administered by the Cohort and Registry Administration (CARA) Core, a Virginia Commonwealth University shared resource. We would like to extend our gratitude to all of the MATR twins who took part in this study.

---

**Supplementary Figure 1.**
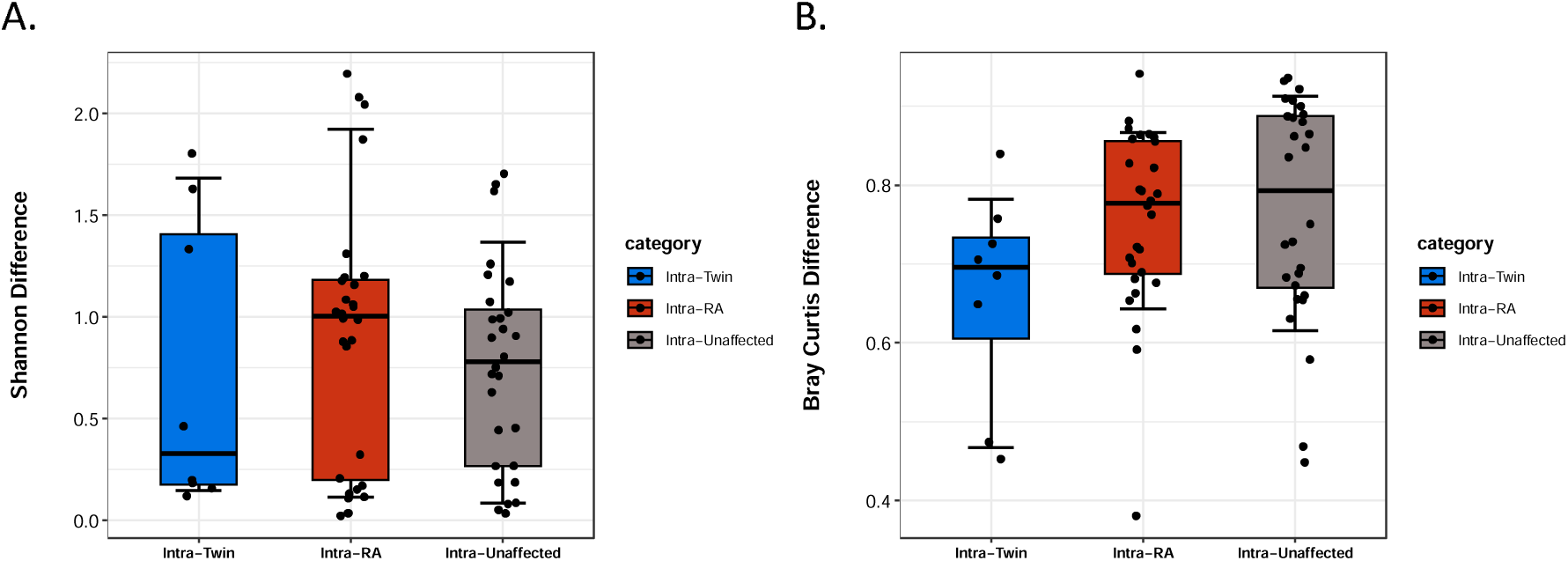
Comparison of gut microbial diversity between individuals of shared genetic background versus shared diagnosis. Distribution of intra-RA, intra-unaffected, and intra-twin differences are shown in red, grey, and blue, respectively for (A) alpha diversity (Shannon Entropy) and (B) beta diversity (Bray-Curtis).

**Supplementary Figure 2.**
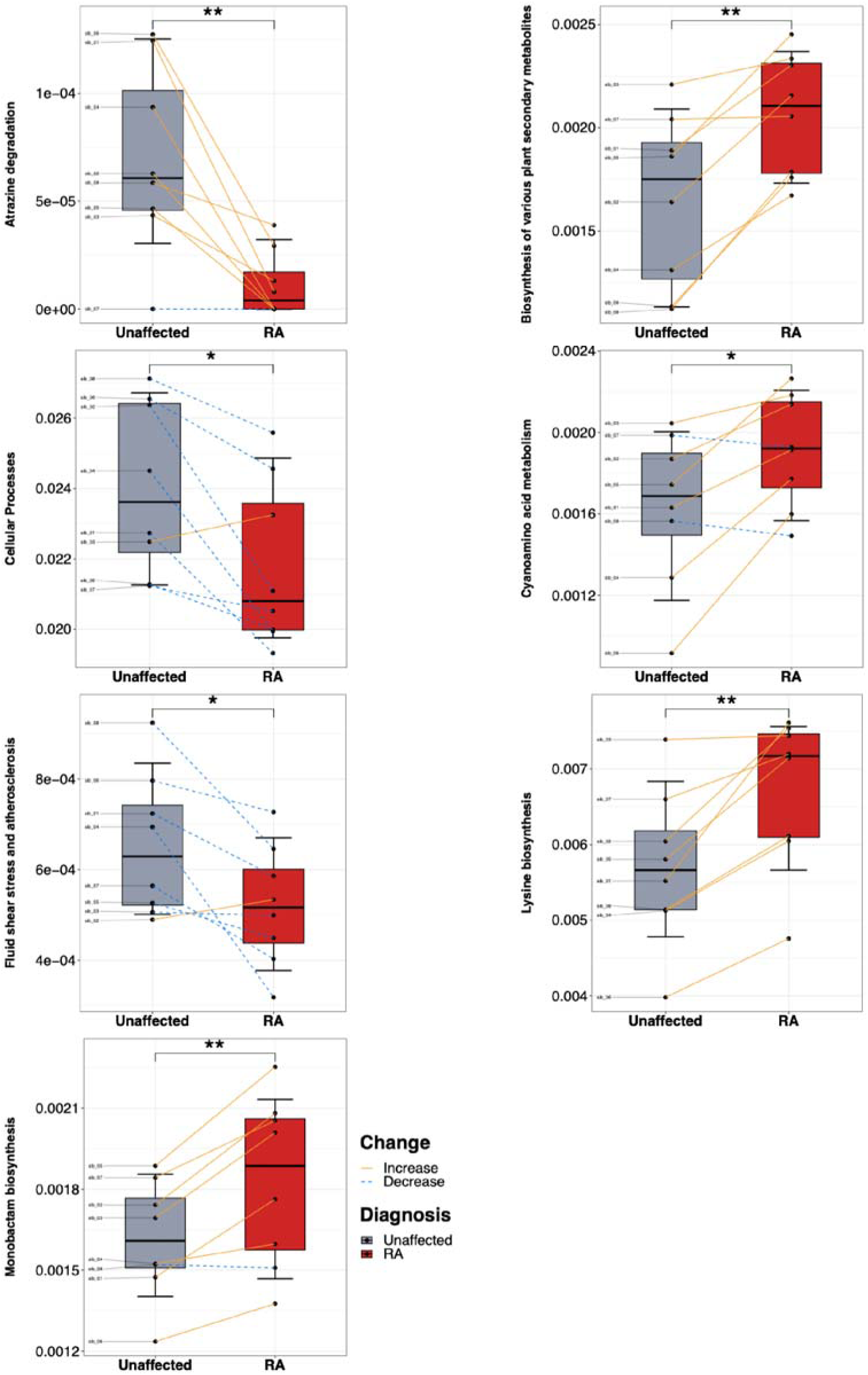
Differential pathways between RA and unaffected twins using the BriteKO hierarchy. Y axis represents relative abundance. *p<0.05, **p<0.01.

**Supplementary Figure 3.**
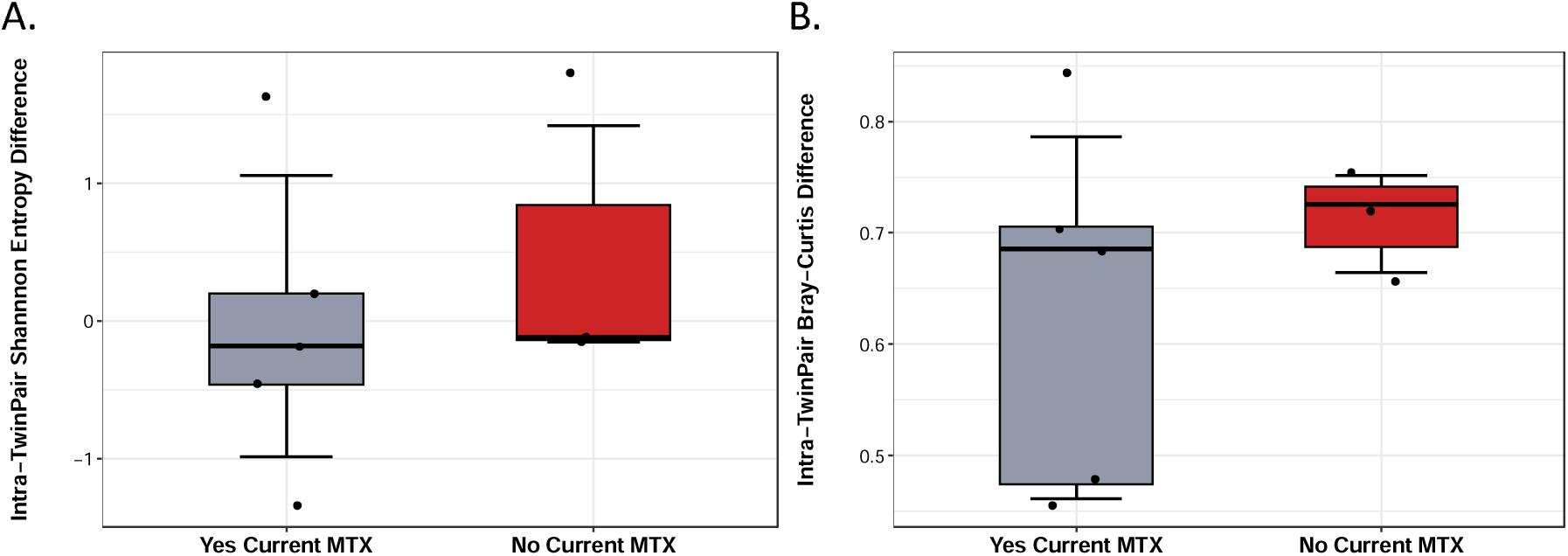
Intra-twin pair microbiome alpha and beta diversity distance as a function of current methotrexate treatment. Distribution of intra-twin pair differences in (A) alpha diversity (Shannon Entropy) and (B) beta diversity (Bray-Curtis) comparing those pairs in which the RA-affected twin is treated with MTX (left) or not (right).

**Supplementary Figure 4.**
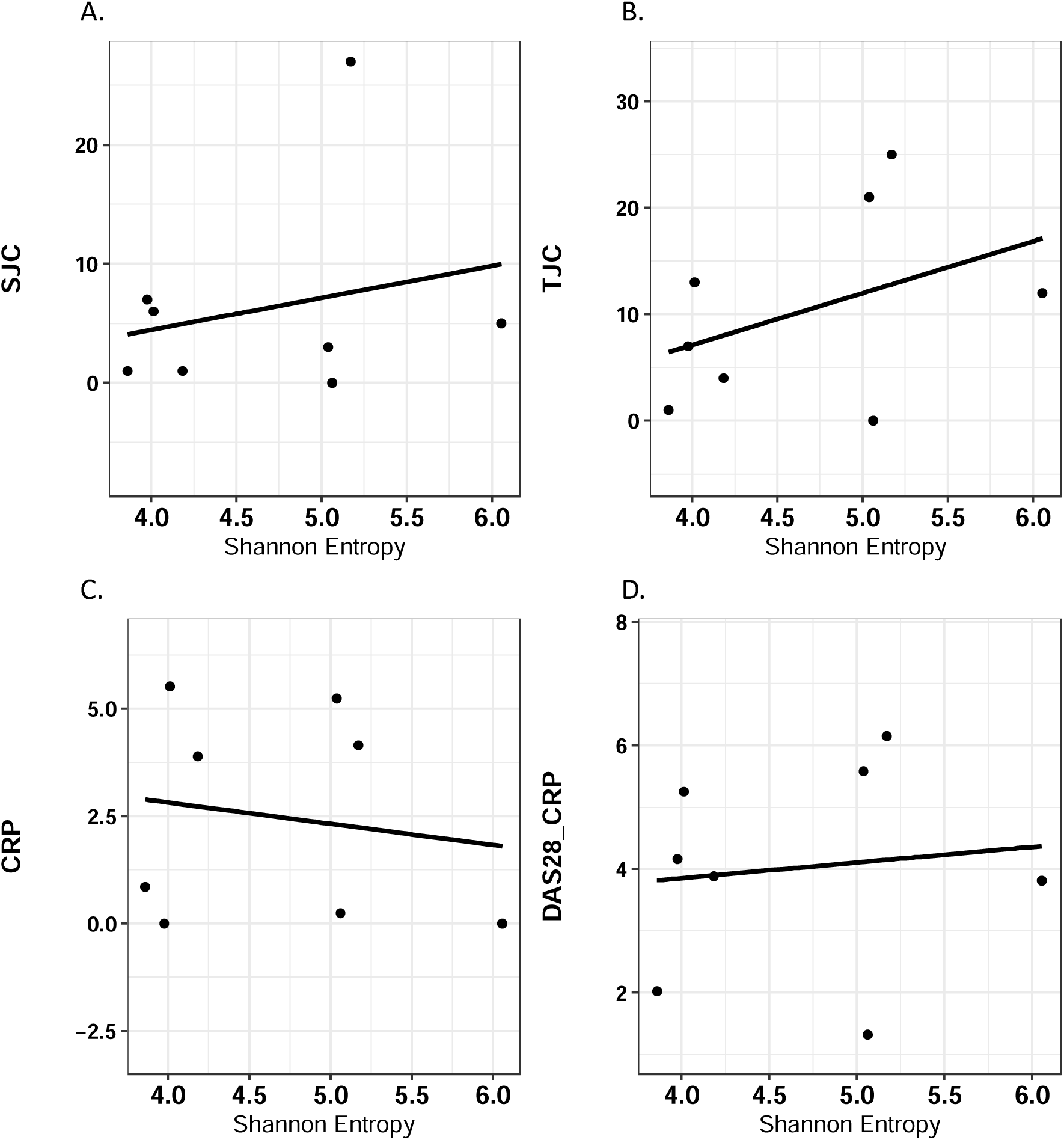
Correlation of alpha diversity (Shannon Entropy) with (A) SJC (rho=0.11, p=0.8), (B) TJC (rho=0.36, p=0.39), (C) CRP (rho=-0.10, p=0.82), and (D) DAS28-CRP (rho=0.12, p=0.78).

**Supplementary Figure 5.**
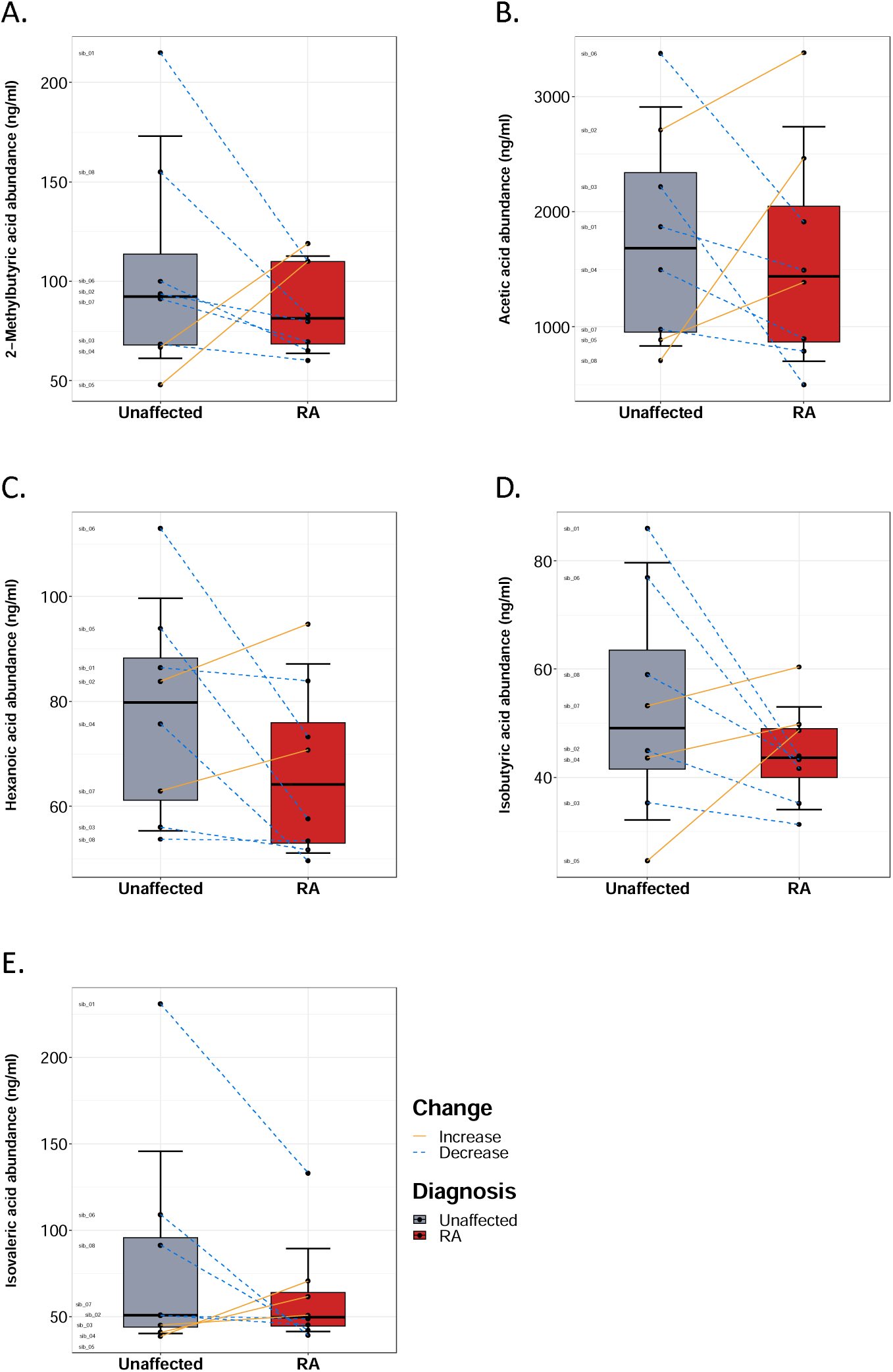
Comparison of serum SCFA concentrations between unaffected and RA twins. Plots are shown for (A) 2-methylbutyric acid, (B) acetic acid (C) hexanoic acid, (D) isobutyric acid, and (E) isovaleric acid. Box plots represent median and interquartile range. Lines are drawn between MZ pairs. P-values calculated with paired t test; *p<0.05.

**Supplementary Figure 6.**
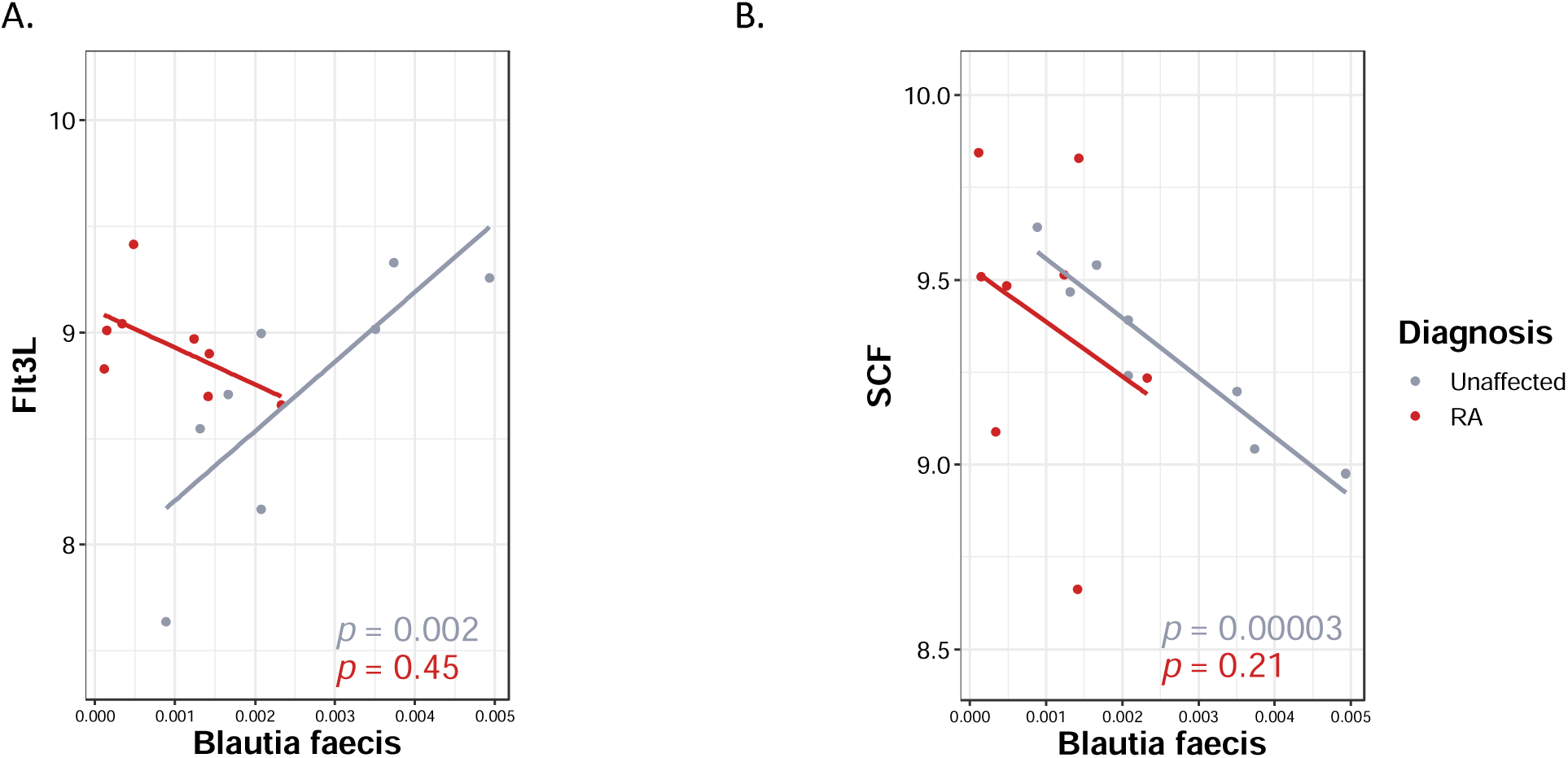
Differential Spearman correlations between RA and unaffected twins involving. B. faecis and cytokines (A) Flt3L and (B) SCF.

**Supplementary Figure 7.**
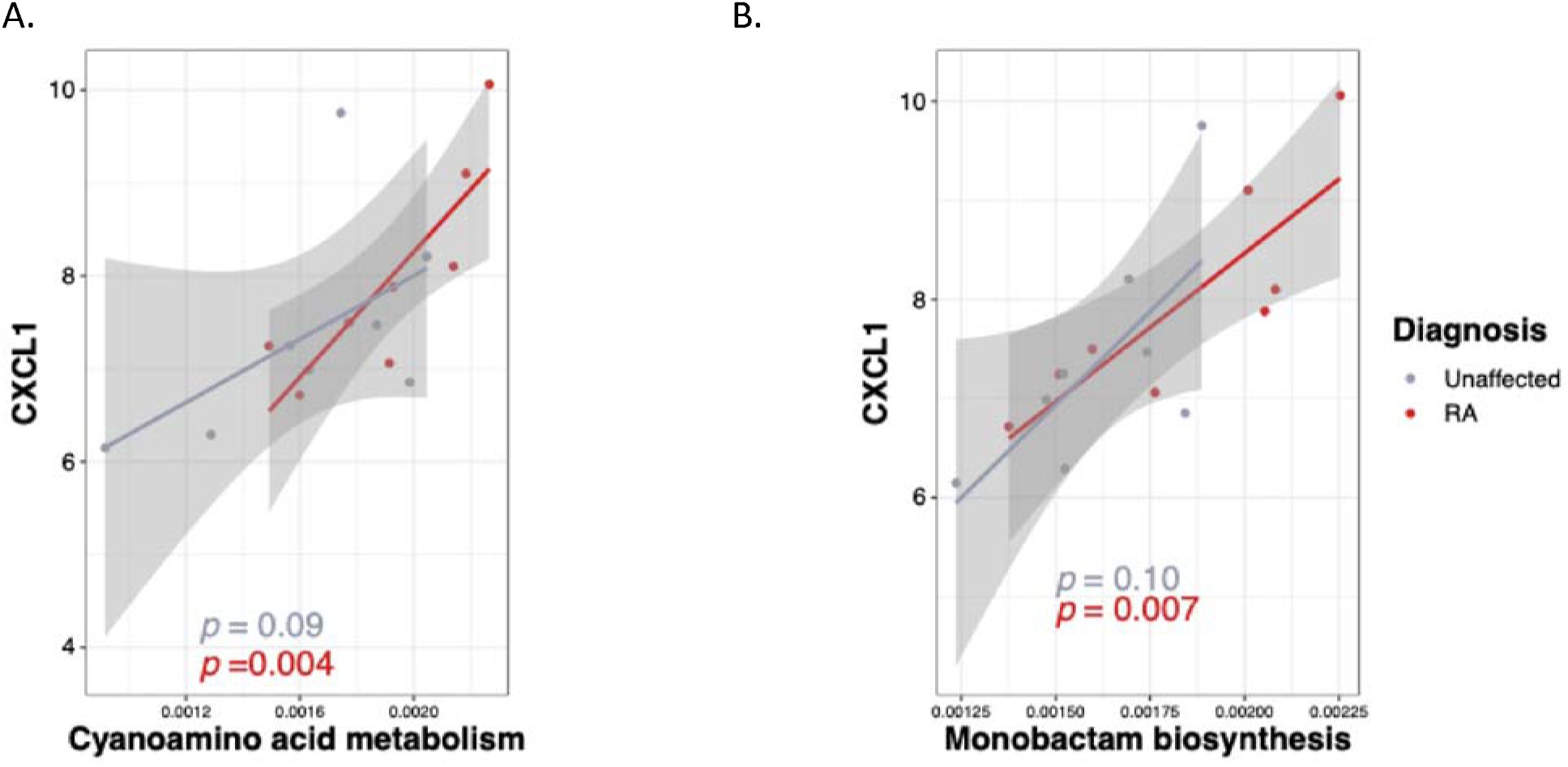
Spearman correlations between differentially expressed pathways (A) Monobactam synthesis and (B) Cyanoamino acid metabolism and cytokine CXCL1 (enriched in RA twins).

**Supplementary Table 1.** List of fecal and plasma autoantibodies analyzed by multiplex autoantibody assay.

| **Fecal autoantibodies** |
| --- |
| Biglycan 247-266 sm-1 cyclic IgG |
| Clusterin 231-250 sm-1 cyclic IgG |
| ApoE 277-296 Cit2 sm2 cyclic IgG |
| H2B/a 62-81 cyclic IgG |
| ApoA1 Cit IgG |
| FibrinogenB 36-52 cit IgG |
| FibrinogenB 246-267 cit IgG |
| Vimentin 58-77 cit3 cyclic small-1 IgG |
| FibrinogenA 41-60 cyclic IgA |
| ApoA1 231-248 IgG |
| Histones2A IgA |
| FibrinogenB 54-72 IgA |
| Fibrinogen IgA |
| FibrinogenA 616-635 small cyclic IgA |
| Filaggrin 48-65 v1 cyclic IgA |
| Vimentin 1-16 IgA |
| FibrinogenA 556-575 sm cyclic IgA |
| Histones2B cit IgA |
| FibrinogenB 246-267 IgG |
| H2A/a-2 1-20 IgA |
| Filaggrin 48-65 arg2 v1 cyclic IgA |
| FibrinogenA 211-230 small cyclic IgA |
| Histones2B IgA |
| Vimentin 1-16 IgG |
| FibrinogenB 246-267 IgA |
| Vimentin 58-77 cyclic small-1 IgA |
| Enolase-1A 5-21 cit IgG |
| Histones2A cit IgG |
| FibrinogenA 582-599 cit IgG |
| H2A/a 1-20 cit sm-2 cyclic IgG |
| Clusterin 231-250 cit sm-1 cyclic IgG |
| FibrinogenA 211-230 cit small cyclic IgA |
| FibrinogenA 582-599 IgA |
| CCP IgA |
| Clusterin 231-250 sm-1 cyclic IgA |
| Vimentin 58-77 cyclic small-1 IgG |
| ApoA1 231-248 cit1 IgG |
| ApoA1 231-248 IgA |
| Filaggrin 48-65 cit2 v1 cyclic IgA |
| FibrinogenB 36-52 cit IgA |
| Vimentin 58-77 cit3 cyclic small-1 IgA |
| ApoE 277-296 Cit2 sm2 cyclic IgA |
| Fibronectin cit 1029-1042 cit2 IgA |
| FibrinogenA 556-575 cit sm cyclic IgG |
| Clusterin 231-250 cit sm-1 cyclic IgA |
| Fibrinogen cit IgA |
| Clusterin 221-240 cit cyclic IgA |
| Biglycan 247-266 sm-1 cyclic IgA |
| FibrinogenB 54-72 cit1 IgA |
| FibrinogenB 246-267 cit IgA |
| H2B/a 62-81 cit cyclic IgA |
| FibrinogenA 27-43 cit IgA |
| FibrinogenA 556-575 cit sm cyclic IgA |
| FibrinogenA 616-635 cit3 small cyclic IgA |
| Filaggrin 48-65 cit2 v1 cyclic IgG |
| Enolase-1A 5-21 IgG |
| ApoA1 231-248 cit1 IgA |
| ApoA1 Cit IgA |
| ApolipoproteinE CIT IgA |
| Vimentin cit IgA |
| FibrinogenA 41-60 cit3 cyclic IgA |
| Biglycan 247-266 cit sm-1 cyclic IgG |
| Histones2A cit IgA |
| FibrinogenA 582-599 cit IgA |
| Clusterin 221-240 cit cyclic IgG |
| FibrinogenA 616-635 small cyclic IgG |
| Enolase-1A 5-21 IgA |
| H2B/a 62-81 cyclic IgA |
| Vimentin cit IgG |
| Enolase-1A 5-21 cit IgA |
| FibrinogenA 41-60 cyclic IgG |
| FibrinogenA 616-635 cit3 small cyclic IgG |
| Biglycan 247-266 cit sm-1 cyclic IgA |
| H2A/a-2 1-20 IgG |
| Vimentin IgA |
| Vimentin 1-16 cit IgA |
| ApoA1 IgA |
| FibrinogenA 582-599 IgG |
| Filaggrin 48-65 v1 cyclic IgG |
| FibrinogenB 54-72 cit1 IgG |
| H2A/a 1-20 cit sm-2 cyclic IgA |
| H2B/a 62-81 cit cyclic IgG |
| FibrinogenA 211-230 small cyclic IgG |
| ApoA1 IgG |
| ApolipoproteinE CIT IgG |
| H2A/a-2 1-20 cit IgG |
| Vimentin 1-16 cit IgG |
| Histones2B IgG |
| FibrinogenB 54-72 IgG |
| Fibronectin cit IgG |
| FibrinogenA 41-60 cit3 cyclic IgG |
| CCP IgG |
| H2A/a-2 1-20 cit IgA |
| Histones2A IgG |
| FibrinogenA 27-43 cit IgG |
| Vimentin IgG |
| Fibrinogen IgG |
| Fibrinogen cit IgG |
| Filaggrin 48-65 arg2 v1 cyclic IgG |
| Fibronectin cit IgA |
| FibrinogenA 556-575 sm cyclic IgG |
| Histones2B cit IgG |
| Fibronectin cit 1029-1042 cit2 IgG |
| FibrinogenA 211-230 cit small cyclic IgG |
| **Plasma autoantibodies** |
| FibrinogenA 41-60 cit3 cyclic IgA |
| FibrinogenA 27-43 cit IgA |
| Biglycan 247-266 cit sm-1 cyclic IgA |
| FibrinogenA 41-60 cyclic IgA |
| H2B/a 62-81 cyclic IgA |
| ApoE 277-296 Cit2 sm2 cyclic IgG |
| Enolase-1A 5-21 cit IgA |
| FibrinogenA 556-575 sm cyclic IgA |
| FibrinogenB 54-72 IgA |
| Vimentin 1-16 IgA |
| CCP IgA |
| Enolase-1A 5-21 cit IgA |
| CCP IgG |
| Filaggrin 48-65 v1 cyclic IgA |
| Biglycan 247-266 sm-1 cyclic IgA |
| FibrinogenA 27-43 cit IgG |
| FibrinogenA 41-60 cyclic IgG |
| FibrinogenA 556-575 sm cyclic IgG |
| FibrinogenA 582-599 cit IgG |
| FibrinogenA 582-599 IgG |
| FibrinogenB 246-267 cit IgG |
| FibrinogenA 616-635 cit3 small cyclic IgG |
| ApoA1 231-248 cit1 IgG |
| FibrinogenA 616-635 small cyclic IgA |
| Clusterin 221-240 cit cyclic IgG |
| Vimentin 1-16 cit IgG |
| FibrinogenB 36-52 cit IgA |
| FibrinogenA 211-230 small cyclic IgA |
| FibrinogenA 41-60 cit3 cyclic IgG |
| FibrinogenB 246-267 IgG |
| Filaggrin 48-65 cit2 v1 cyclic IgA |
| Clusterin 231-250 sm-1 cyclic IgA |
| FibrinogenA 616-635 small cyclic IgG |
| FibrinogenB 36-52 cit IgG |
| Clusterin 231-250 sm-1 cyclic IgG |
| H2B/a 62-81 cyclic IgG |
| FibrinogenA 582-599 cit IgA |
| Fibronectin cit 1029-1042 cit2 IgA |
| Vimentin cit IgG |
| FibrinogenA 211-230 cit small cyclic IgG |
| Clusterin 221-240 cit cyclic IgA |
| Biglycan 247-266 cit sm-1 cyclic IgG |
| ApoA1 231-248 IgA |
| Vimentin 1-16 IgG |
| ApoA1 231-248 cit1 IgA |
| ApoA1 231-248 IgG |
| ApoE 277-296 Cit2 sm2 cyclic IgA |
| Filaggrin 48-65 cit2 v1 cyclic IgG |
| FibrinogenA 616-635 cit3 small cyclic IgA |
| FibrinogenA 582-599 IgA |
| H2A/a 1-20 cit sm-2 cyclic IgG |
| Enolase-1A 5-21 IgA |
| FibrinogenB 54-72 cit1 IgG |
| Fibrinogen IgA |
| FibrinogenA 211-230 small cyclic IgG |
| FibrinogenB 54-72 IgG |
| Filaggrin 48-65 v1 cyclic IgG |
| FibrinogenB 246-267 IgA |
| Vimentin 58-77 cyclic small-1 IgG |
| Fibronectin cit 1029-1042 cit2 IgG |
| FibrinogenB 54-72 cit1 IgA |
| Histones2A IgG |
| Biglycan 247-266 sm-1 cyclic IgG |
| Filaggrin 48-65 arg2 v1 cyclic IgG |
| Filaggrin 48-65 arg2 v1 cyclic IgA |
| FibrinogenA 556-575 cit sm cyclic IgG |
| FibrinogenB 246-267 cit IgA |
| Clusterin 231-250 cit sm-1 cyclic IgG |
| Vimentin 58-77 cyclic small-1 IgA |
| H2A/a-2 1-20 cit IgG |
| Enolase-1A 5-21 IgG |
| H2A/a-2 1-20 cit IgA |
| Histones2A IgA |
| Histones2B cit IgG |
| ApolipoproteinE CIT IgA |
| Fibrinogen cit IgA |
| Histones2B cit IgA |
| FibrinogenA 211-230 cit small cyclic IgA |
| Vimentin IgA |
| Fibrinogen cit IgG |
| Vimentin 58-77 cit3 cyclic small-1 IgG |
| Fibronectin cit IgG |
| ApoA1 Cit IgG |
| ApoA1 IgG |
| H2A/a-2 1-20 IgA |
| Fibronectin cit IgA |
| ApolipoproteinE CIT IgG |
| Fibrinogen IgG |
| H2A/a-2 1-20 IgG |
| Clusterin 231-250 cit sm-1 cyclic IgA |
| H2B/a 62-81 cit cyclic IgA |
| Histones2B IgA |
| Vimentin 1-16 cit IgA |
| Vimentin IgG |
| Vimentin cit IgA |
| ApoA1 Cit IgA |
| H2B/a 62-81 cit cyclic IgG |
| Histones2B IgG |
| ApoA1 IgA |
| Histones2A cit IgG |
| H2A/a 1-20 cit sm-2 cyclic IgA |
| Histones2A cit IgA |
| FibrinogenA 556-575 cit sm cyclic IgA |
| Vimentin 58-77 cit3 cyclic small-1 IgA |

**Supplementary Table 2.** Elevated autoantibodies in RA twins.

| Autoantibody level (MFI) | Unaffected | RA | p-value |
| --- | --- | --- | --- |
| **Fecal – mean (SD)** |  |  |  |
| Clusterin 231-250 sm-1 cyclic IgG | 15.0 (2.1) | 16.6 (2.2) | 0.040 |
| **Plasma – mean (SD)** |  |  |  |
| FibrinogenA 556-575 sm cyclic IgA | 34.8 (13.4) | 46.6 (16.8) | 0.019 |
| Biglycan 247-266 cit sm-1 cyclic IgA | 26.1 (9.3) | 37.3 (11.1) | 0.020 |
| H2B-a 62-81 cyclic IgA | 33.3 (11.1) | 45.8 (16.7) | 0.021 |
| FibrinogenA 41-60 cyclic IgA | 29.2 (11.5) | 41.4 (15.0) | 0.023 |
| FibrinogenA 27-43 cit IgA | 23.3 (6.1) | 32.5 (7.1) | 0.026 |
| FibrinogenA 41-60 cit3 cyclic IgA | 21.6 (9.2) | 33.9 (13.8) | 0.027 |
| FibrinogenA 582-599 IgA | 29.7 (8.4) | 40.1 (13.6) | 0.030 |
| ApoE 277-296 Cit2 sm2 cyclic IgG | 69.7 (11.1) | 86.1 (16.6) | 0.031 |
| Vimentin 1-16 cit IgA | 54.6 (20.4) | 69.0 (18.3) | 0.032 |
| Enolase-1A 5-21 cit IgA | 60.1 (14.6) | 186.3 (138.7) | 0.035 |
| Enolase-1A 5-21 cit IgG | 115.3 (113.9) | 2880.6 (3022.1) | 0.042 |
| ApoA1 231-248 IgA | 20.3 (4.7) | 26.1 (6.3) | 0.047 |
| CCP IgA | 128.6 (262.1) | 442.6 (460.8) | 0.047 |
| FibrinogenB 36-52 cit IgA | 35.3 (11.1) | 46.3 (11.4) | 0.047 |

